# Atypical somatic symptoms in adults with prolonged recovery from mild traumatic brain injury

**DOI:** 10.1101/19004622

**Authors:** Jacob L. Stubbs, Katherine E. Green, Noah D. Silverberg, Andrew Howard, Amrit Dhariwal, Jeffrey R. Brubacher, Naisan Garraway, Manraj K. S. Heran, Mypinder S. Sekhon, Angela Aquino, Victoria Purcell, James S. Hutchison, Ivan J. Torres, William J. Panenka, on behalf of the National biobank and database of patients with traumatic brain injury (CanTBI) investigators and the Canadian Traumatic brain injury Research Consortium (CTRC)

## Abstract

Somatization may contribute to persistent symptoms after mild traumatic brain injury (mTBI). In two independently-recruited study samples, we characterized the extent to which symptoms atypical of mTBI but typical for somatoform disorders (e.g., gastrointestinal upset, joint pain) were present in adult patients with prolonged recovery following mTBI. The first sample was cross-sectional and consisted of mTBI patients recruited from the community who reported ongoing symptoms attributable to a previous mTBI (*n* = 16) along with a healthy control group (*n* = 15). The second sample consisted of patients with mTBI prospectively recruited from a Level 1 trauma center who had either good recovery (GOSE = 8; n = 33) or poor recovery (GOSE < 8; n = 29). In all participants, we evaluated atypical somatic symptoms using the Patient Health Questionnaire-15 and typical post-concussion symptoms with the Rivermead Post-Concussion Symptom Questionnaire. Participants with poor recovery from mTBI had significantly higher ‘atypical’ somatic symptoms as compared to the healthy control group in Sample 1 (*b* = 4.308, *p* = 9.43E^-5^) and to mTBI patients with good recovery in Sample 2 (*b* = 3.287, *p* = 6.83E^-04^). As would be expected, participants with poor outcome in Sample 2 had a higher burden of typical rather than atypical symptoms (*t*(28) = 3.675, *p* = 9.97E^-04^, *d* = 0.94). However, participants with poor recovery still reported atypical somatic symptoms that were significantly higher (1.4 standard deviations, on average) than those with good recovery. Our results suggest that although ‘typical’ post-concussion symptoms predominate after mTBI, a broad range of somatic symptoms also frequently accompanies mTBI, and that somatization may represent an important, modifiable factor in mTBI recovery.

## INTRODUCTION

An estimated forty-two million people experience mild traumatic brain injuries (mTBI) worldwide annually.^1^ Symptoms generally resolve within the first week; however, a substantial number of patients experience chronic symptoms for months or years after injury, leading to significant disability and functional impairment.^2, 3^ Although there are many factors that influence the recovery trajectory, pre- and post-injury mental health problems are the strongest established contributor to poor recovery and functional limitation after mTBI.^4, 5^

The term post-concussion syndrome (PCS) dates back to at least World War II where, based mainly on studies of soldiers with blast injury (i.e., ‘shell shock’), it was characterized by headache, dizziness, fatigue, tinnitus, memory impairment, poor concentration and nervousness.^6^ The Rivermead Post-Concussion Syndrome questionnaire (RPQ) was developed in 1995 by aggregating the 16 most commonly reported post-concussion symptoms,^7^ and remains endorsed by the National Institute for Neurological Diseases and Stroke Common Data Elements as the instrument of choice for evaluating post-concussion symptoms in adults. Although there is significant ongoing debate as to the etiology of some of the symptoms the endurance of this legacy instrument, unmodified, reflects at least some consensus that these are the cardinal features expected after a brain injury.

Somatization is a process whereby psychological distress manifests as physical symptoms, which can occur in the presence or absence of organic pathology.^8^ Historically ‘medically unexplained symptoms’ have served as the foundation for the diagnosis of somatization and somatoform disorders, recognizing that, at times, it can be difficult or impossible to differentiate these symptoms from those resulting from general medical conditions (GMC).^9^ When these symptoms do occur in the context of an associated GMC, somatization can be diagnosed when the severity of the symptom exceeds what can be logically attributed to the GMC. When somatization is left undiagnosed and untreated, these physical symptoms and associated dysfunction typically persist or worsen, which leads to considerable costs to society and the health care system.^10^ In addition, patients are often at risk for iatrogenic harm from unnecessary medical investigations and treatments.^11^

There is an emerging literature pointing to an etiological role for somatization in prolonging the recovery process after mTBI.^12-17^ Two previous studies in pediatric patients recruited from emergency departments have examined measures of somatization after mTBI, both finding that higher measures of somatization were associated with prolonged symptom duration.^12, 17^ A recent study of high school and collegiate athletes found pre-injury somatic symptom scores to be the strongest pre-morbid predictor of post-concussive symptom duration.^13^ However, like most other studies analyzing somatic symptoms after mTBI, Nelson et al. (2016) evaluated somatization using a composite score reflective of somatic complaints across multiple body systems, and did not distinguish the somatic symptoms that would be conventionally associated with mTBI (e.g., headache and dizziness) from others that would be more likely related to somatoform pathology (e.g., intestinal upset, diffuse body pains, etc). In so doing, they are potentially conflating organic brain injury with psychopathology.

Three studies, performed in the context of comprehensive health assessments in military personnel, have used somatic symptom scales broken down by item to evaluate somatic symptoms post-TBI, allowing for an assessment of the type of somatic symptoms experienced after mTBI. These studies consistently document an elevated level of somatic symptoms not plausibly related to head injury after TBI (e.g., chest pain, heart pounding or racing, shortness of breath).^14-16^ Critically, these atypical somatic symptoms may be prognostic, as a study in military personnel by Lee et al. (2015) found that an aggregated metric of pre-injury somatic symptoms was associated with the subsequent development of post-concussion syndrome.^16^ However, while these military studies suggest significantly heightened somatic symptoms post-TBI, the high prevalence of psychiatric comorbidities confounds causative inference. For example, Hoge et al. (2008) documented a 44% prevalence of post-traumatic stress disorder (PTSD) and 27% prevalence of major depression after mTBI with loss of consciousness, and concluded that PTSD and depression are strongly associated with physical health problems upon return from deployment.^15^ They further suggest that PTSD or depression mediate the majority of the relationship between mTBI and subsequent somatic complaints.^15^ If this is correct, given lower rates of PTSD and depression in the civilian population as compared to military members,^18^ after civilian mTBI we might expect lower levels of somatization than in a military sample. However, if elevated somatization contributes to poorer recovery from mTBI independent of other mental health concerns, then rates in civilians with persistent symptoms might also be high.

Our aim was to evaluate symptoms *atypical* of mTBI (i.e., symptoms not typically related to the mechanism of injury) in adult civilians who had poor recovery from their mTBI and who had no other pre-injury history of psychopathology. First in an initial pilot study, and subsequently replicated in a prospectively-recruited sample, we administered a modified version of the most widely used assessment instrument for somatization symptoms, the Patient Health Questionnaire (PHQ-15), that had the four questions that reflect typical post-concussion complaints (i.e., headache, dizziness, insomnia and fatigue) removed. We hypothesized that mTBI patients with poor recovery would report a higher severity of symptoms not typically associated with brain injury (e.g., gastrointestinal upset, sexual dysfunction, etc.) compared to those with good recovery and also as compared to a healthy control group. Support for this hypothesis would provide further evidence for an association between somatization and prolonged recovery from mTBI.

## MATERIALS AND METHODS

This study occurred in two phases and drew from two independently-recruited sources (recruitment flow chart shown in Figure 1). Informed consent was provided by all participants, and studies were approved by the University of British Columbia (UBC) Clinical Research Ethics Board (H16-01307 and H15-01063).

**Figure 1.**
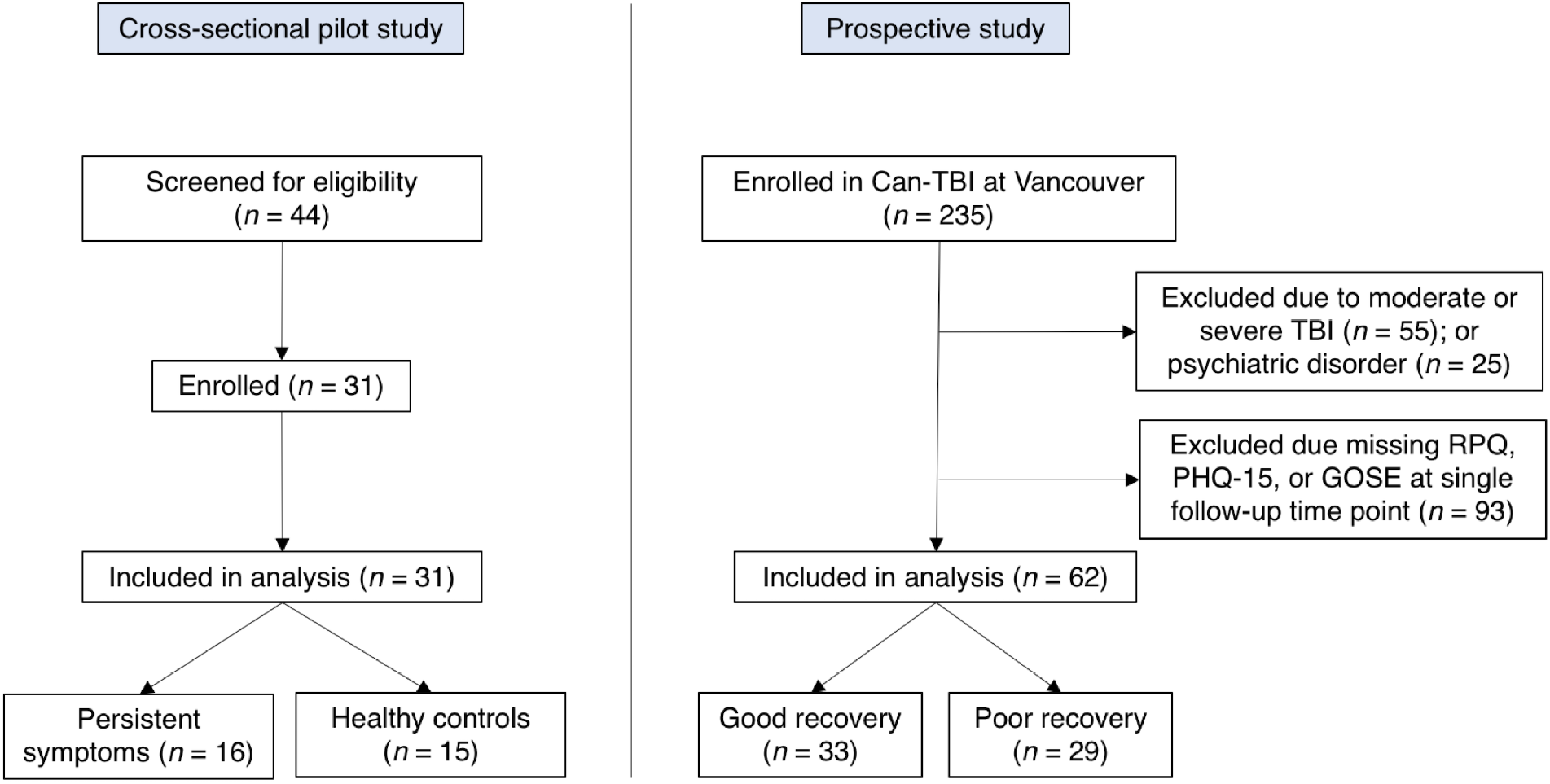
Flow chart outlining participant recruitment for both study samples.

### Participants and study design: Sample 1

Initially, in the context of an exploratory pilot study, we recruited a cross-sectional sample of 16 patients who had sustained an mTBI more than 30 days previously and who self-reported persistent symptoms from that mTBI. We also recruited 15 healthy controls from the community through an institutional newsletter. Healthy controls were included if they reported no TBI during their lifetime. Mild traumatic brain injury was assessed with the Ohio State University TBI identification method in accordance with the World Health Organization definition of mTBI.^19, 20^ Mild traumatic brain injury participants and healthy controls were had to be between 18 and 50 years of age and fluent in English, and were excluded if they self-reported any diagnosed psychiatric illness or substance abuse.

### Participants and study design: Sample 2

Based on results from our initial pilot study we then assessed an additional subset of participants from an ongoing prospectively observational study of TBI patients entitled ‘A national biobank and database for patients with TBI (CanTBI).’ Participants were included in the broader CanTBI study if they (i) had a diagnosis of a mild, moderate, or severe TBI made by a physician; (ii) had at least one blood draw for research purposes within 24 hours of injury; and (iii) were fluent in English or French. Participants were excluded from CanTBI if they (i) had any neurodevelopmental or ongoing neurological disorder; (ii) had suffered a stroke, cardiac arrest, or had significant disruptive neurological issues; (iii) were brain dead or suffered from a terminal illness (life expectancy < 12 months at assessment); (iv) or were currently a prisoner, patient in custody, or enrolled in an intervention trial. From this broader CanTBI study, we evaluated somatic symptom scores in adult patients with mTBI who had no other diagnosed psychiatric illness or substance abuse (Figure 1). Participants were excluded from our analysis if they had (i) sustained a moderate or severe TBI; (ii) were less than 18 years of age; (iii) had not completed both the Rivermead and the Patient Health Questionnaire-15 at a follow-up time point three months post-TBI or greater; and (iv) had a history of diagnosed psychiatric illness. CanTBI participants were classified into either a ‘good recovery’ group or ‘poor recovery’ group based on the Glasgow Outcome Scale Extended (operationalized below).

### Measures

In both study samples, mTBI symptoms were assessed with the Rivermead Post-Concussion Symptoms Questionnaire (RPQ).^7^ The RPQ consists of 16 questions about post-concussion symptoms on a Likert scale ranging from 0 (‘not experienced at all’) to 4 (‘a severe problem’). All scores of 1 or greater were included in total score calculations, for a potential maximum score of 64. In both study samples, somatic symptoms experienced in the 4 weeks preceding evaluation were measured using the Patient Health Questionnaire (PHQ-15), which is a 15-question subset of the full PHQ.^21^ The PHQ-15 is a commonly used instrument for the assessment of somatic symptoms that is a both valid and reliable proxy measure of somatization.^22, 23^ It is used to assess 15 non-specific physical symptoms spanning multiple organ systems.^21^ PHQ-15 scores possess moderate-to-good diagnostic accuracy for identifying somatoform disorder as assessed with a structured interview for the Diagnostic and Statistics Manual (DSM)-IV,^24^ and somatic symptom disorder assessed with a structured interview for the DSM-V.^25^ Each PHQ-15 item can be rated as ‘not bothered at all’, ‘bothered a little’, or ‘bothered a lot’, resulting in a score of 0, 1, or 2 points per question respectively, for a range from 0 – 30. A score of 1 or more on a PHQ-15 item was considered a positive endorsement of that somatic symptom. All data were collected with the secure, electronic REDCap Data Capture Tool hosted at the BC Children’s Hospital Research Institute.^26^

To examine symptoms atypical of mTBI in both study samples, we excluded PHQ-15 items *a priori* that were most likely etiologically related to mTBI (a method previously employed by Lee et al. (2015)). Specifically, we excluded the questions about headaches, dizziness, feeling tired or having low energy, and trouble sleeping. The remaining eleven PHQ-15 items were considered ‘atypical’ for mTBI, and included stomach pain, back pain, pain in arms, legs or joints, menstrual cramps, chest pain, fainting spells, heart pounding or racing, shortness of breath, problems during intercourse, constipation, loose bowels or diarrhea, or nausea, bloating, or indigestion. Where listed, ‘PHQ-15’ is the total score on the full PHQ-15 (maximum score 30 and including all 15 questions) and the ‘atypical’ symptom subset is the total score for the 11-question subset of the PHQ-15 which queries only the symptoms that would be considered ‘atypical’ after mTBI (maximum score 22).

In the prospectively-recruited sample, we used the Glasgow Outcome Scale Extended (GOSE) to evaluate outcome from mTBI.^27^ The GOSE has eight categories to measure global neurological function or death; it is a sensitive outcome measures across the injury severity spectrum, including in mTBI.^28, 29^ It parallels other indicators of recovery including post-concussion symptoms,^30^ and is endorsed as one of the few core mTBI outcome measure by the NINDS Common Data Elements group.^31^ As in other large multi-site mTBI studies^32^ participants with a GOSE score of 8/8 were considered to have ‘good recovery’, while participants with a GOSE score < 8 were considered to have ‘poor recovery’. Of the 62 participants 33 had good recovery, and 29 had poor recovery at the time of assessment.

### Statistical analysis

For between-group comparisons we used independent-samples *t*-tests for continuous variables if normally distributed (as assessed with a Shapiro-Wilk test) or Mann-Whitney *U*-tests for non-normally distributed continuous variables, and Chi-squared tests for categorical variables or Fisher’s Exact Tests for categorical variables if the expected cell count was less than five. To test for differences in RPQ, PHQ-15 total score, and atypical symptom scores from the PHQ-15, we used multiple linear regression models (with *R*^2^ as the measure of effect size), adjusting for age and gender in the cross-sectionally recruited sample, and adjusting for age, gender, and number of days post-injury in the prospectively recruited sample. In the prospectively-recruited sample, we assessed the relative symptom burden of typical mTBI symptoms (RPQ), global somatic symptoms (PHQ-15 total score), and atypical somatic symptoms from the PHQ-15 in the group with poor recovery. To do this, we first internally standardized participant scores on each measure into z-scores, using the good recovery group as the reference. We then compared mean z-scores on each of the three outcomes within the poor recovery group using paired *t*-tests with Cohen’s *d* to calculate effect size. This allowed us to determine the predominant symptom burden reported by individuals with poor recovery after mTBI. Statistical analysis was performed using *R* version 3.6.0.^33^

## RESULTS

Demographic and injury-related data, as well as unadjusted RPQ and PHQ-15 scores for both study samples are presented in Table 1. In the cross-sectionally recruited sample, there were no significant differences in age or gender between the symptomatic and healthy control groups. There were no significant differences in age and gender between the good and poor recovery groups in the prospectively recruited study, nor were there differences in peri-injury variables including LOC, GCS, mechanism of injury, whether or not participants received a head CT scan or had acute trauma-related finding on those CT scans. The cross-sectionally recruited symptomatic group and the prospectively recruited poor recovery group were not significantly different in the number of days post-injury (*U* = 209, *p* = 0.843), nor were the prospectively-recruited good and poor recovery groups (*U* = 424.5, *p* = 0.450).

**Table 1.** Demographic, injury, and outcome metrics for the both study samples.

|  | Study 1 (cross-sectional community) |  |  | Study 2 (prospectively recruited from ER) |  |  |
| --- | --- | --- | --- | --- | --- | --- |
| | Healthy controls ($n = 15$) | Symptomatic mTBI ($n = 16$) | $p$-value | Good recovery ($n = 33$) | Poor recovery ($n = 29$) | $p$-value |
| Age, mean (SD) | 31.1 (8.0) | 31.6 (6.4) | 0.862 | 40.2 (17.5) | 45.1 (16.1) | 0.139 |
| % female | 80 | 81.3 | $> 0.99$ | 30.3 | 41.4 | 0.52 |
| Years of education, mean (SD)<sup>1</sup> | N/A | N/A | N/A | 16.6 (3.7) | 15.7 ( 3.4) | 0.553 |
| Months post-TBI, mean (SD) | N/A | 5.6 (3.6) | N/A | 8.2 (5.4) | 6.5 (4.7) | 0.45 |
| Cause of injury |  |  |  |  |  |  |
| MVA, $n$ | N/A | 5 | N/A | 12 | 11 | $> 0.99$ |
| Sport, $n$ | N/A | 7 | N/A | 7 | 4 | 0.667 |
| Fall, $n$ | N/A | 3 | N/A | 9 | 7 | $> 0.99$ |
| Other, $n$ | N/A | 1 | N/A | 5 | 7 | 0.375 |
| LOC after injury<sup>2</sup> |  |  |  |  |  |  |
| Yes, $n$ | N/A | 2 | N/A | 13 | 6 | 0.131 |
| No, $n$ | N/A | 11 | N/A | 12 | 15 | 0.414 |
| Suspected or unknown, *n* | N/A | 2 | N/A | 8 | 8 | 0.46 |
| GCS (best prehospital)<sup>3</sup> | N/A | N/A | N/A |  |  |  |
| 15, *n* | N/A | N/A | N/A | 13 | 14 | 0.288 |
| 13-14, *n* | N/A | N/A | N/A | 7 | 3 | 0.288 |
| CT scan |  |  |  |  |  |  |
| Performed, *n* | N/A | N/A | N/A | 22 | 19 | 0.878 |
| Acute findings, *n* | N/A | N/A | N/A | 13 | 6 | 0.148 |
| RPQ, mean (SD) | 2.3 (3.5) | 34.1 (11.3) | 2.90E-06 | 2.9 (5.3) | 18.1 (14.1) | 1.18E-08 |
| PHQ-15, mean (SD) | 2.1 (1.8) | 10.9 (4.2) | 4.66E-06 | 2.6 (2.8) | 8.2 (6.1) | 8.19E-06 |
| <sup>1</sup> Years of education data not available for eight participants; <sup>2</sup> LOC data not available for eight participants;<br><sup>3</sup> GCS data not available for 25 participants |  |  |  |  |  |  |

In the cross-sectional study, as anticipated, post-concussion symptom scores (*b* = 31.650, *p* = 2.27E^-10^, adjusted *R*^2^ = 0.77) and global somatic symptom scores (*b* = 8.757, *p* = 9.57E^-08^, adjusted *R*^2^ = 0.64) were higher in the symptomatic group as compared to the control group, adjusting for age and gender. Our hypothesis was initially supported in this pilot study, as the group with persistent symptoms from their mTBI had significantly higher atypical somatic symptoms as compared to healthy controls adjusting for age and gender (Fig 2; *b* = 4.308, *p* = 9.43E^-5^, adjusted *R*^2^ = 0.39). We then replicated our findings in the prospectively recruited sample: The poor recovery group had significantly higher post-concussion symptom scores (*b* = 16.802, *p* = 6.41E^-07^, adjusted *R*^2^ = 0.33) and global somatic symptom scores (*b* = 5.686, *p* = 2.08E^-05^, adjusted *R*^2^ = 0.25), adjusting for age, gender, and time since injury. Our hypothesis was replicated and again supported in the prospectively recruited sample, with the poor recovery group endorsing significantly higher atypical somatic symptoms than the good recovery group adjusting for age, gender, and time since injury (Fig 2; *b* = 3.287, *p* = 6.83E^-04^, adjusted *R*^2^ = 0.18).

**Figure 2.**
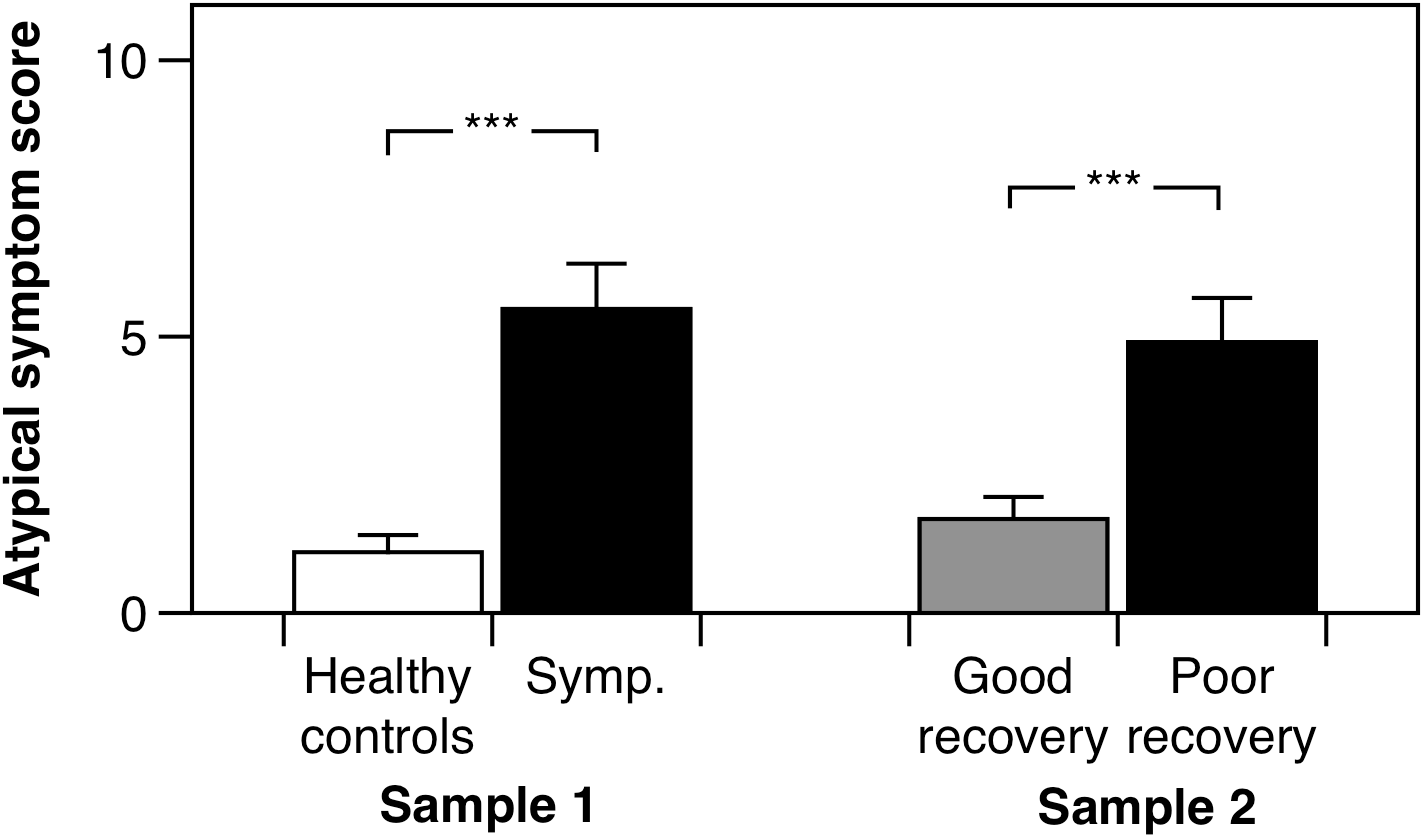
Atypical somatic symptom scores for the cross-sectionally recruited sample (Sample 1) and the prospectively recruited sample (Sample 2). ‘Symp.’ is the subjectively symptomatic group in the cross-sectionally recruited sample. Error bars denote one standard error, and ‘***’ denotes a *p* < 0.001.

We then sought to determine the relative burden of symptom subtypes experienced by those with poor recovery in the prospective sample. Participants with poor recovery endorsed typical post-concussive symptoms (RPQ) 2.9 (SD = 2.7) standard deviations higher, on average, than those with good recovery, global somatic symptoms (PHQ-15) approximately 2.0 (SD = 2.1) standard deviations higher than those with good recovery, and atypical somatic symptoms 1.4 (SD = 1.9) standard deviations higher than the group with good recovery. Using paired t-tests, we found that participants with poor outcome from mTBI had a higher burden of typical post-concussive symptoms than global somatic symptoms (*t*(28) = 3.675, *p* = 9.97E^-04^, *d* = 0.94) and atypical symptoms (*t*(28) = 4.748, *p* = 5.530E^-05^, *d* = 0.99), Figure 3.

**Figure 3.**
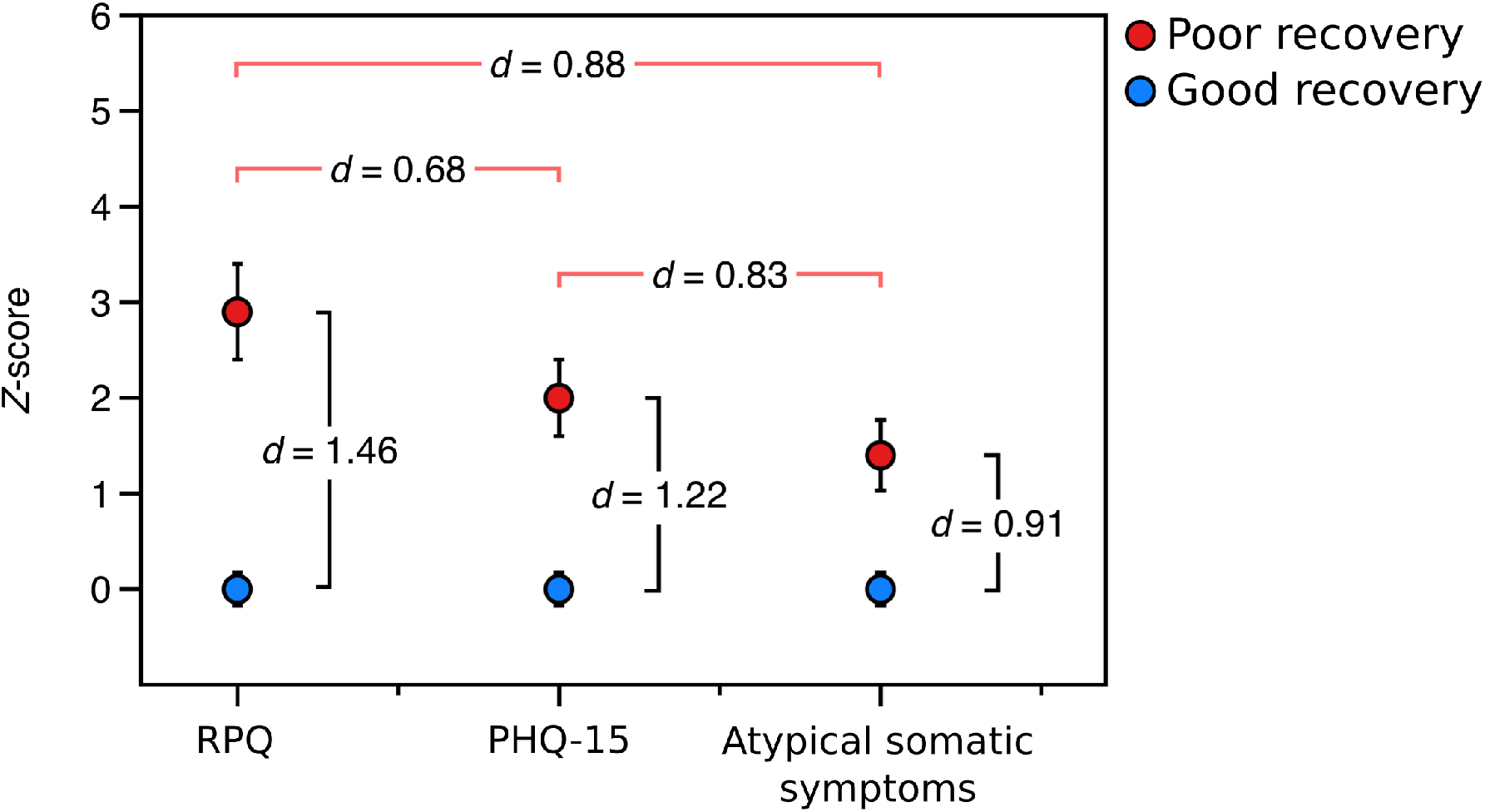
Relative symptom burden in the poor recovery group relative (red) to the good recovery group (blue) in the prospectively-recruited sample (Sample 2). *d*-values between red brackets are values of Cohen’s *d* from paired *t*-tests comparing the relative symptom burden in the group with poor recovery, and *d*-values between black brackets are values of Cohen’s *d* from independent-samples *t*-tests comparing the symptom burden in the poor recovery group to that of the good recovery group. ‘RPQ’ is the Rivermead Post-concussion Questionnaire, ‘PHQ-15’ is the Patient Health Questionnaire-15, and ‘atypical somatic symptoms’ are a subset of questions from the PHQ-15 that exclude those symptoms most plausibly related to mTBI.

## DISCUSSION

Corroborating the limited prior work in this area, we found that heightened post-concussive and global somatic symptoms were associated with prolonged recovery following mTBI. Additionally, we provide evidence that civilians with poor recovery from mTBI experience a significantly greater degree of somatic symptoms which are *atypical* following mTBI, as compared to healthy controls and those with good recovery from mTBI. These results provide further evidence for a role of somatization in persistent symptomology following mTBI.

The only prospective civilian study to link somatization to prolonged recovery from mTBI was recently reported by Nelson and colleagues (2016). Although they did not specifically examine ‘atypical’ symptoms, they did demonstrate a pronounced effect of pre-injury somatization on post-mTBI recovery in athletes. In a univariate analysis on the Brief Symptom Inventory-18,^34^ somatization scores were the strongest pre-injury predictor of recovery duration, even when considered alongside a comprehensive list of pre-injury demographic and history variables (i.e., gender, education, learning disabilities, headache history, number of prior concussions or type and duration of sporting history), psychiatric symptoms (depression, anxiety), cognitive performance, and balance scores. Path analysis indicated that these somatization symptoms likely affected recovery through a mediating effect on post-concussion symptoms, and the authors therefore conclude that somatization may heighten the experience of post-concussion symptoms or increase symptom reporting, subsequently leading to prolonged recovery.

Our study expands on the work of Nelson et al. (2016) by highlighting that not only typical somatic symptoms, but somatic symptoms etiologically unrelated to mTBI, are associated with poor outcome after adult civilian mTBI. This raises the possibility that somatization may be a potentially important modifying factor in the recovery trajectory, and emphasizes the clinical need for measurement of a broad array of somatic symptoms following mTBI. Specifically evaluating ‘atypical’ somatic symptoms may also help to identify individuals suffering from a somatoform disorder that is primarily responsible for, or significantly magnifying their persistent symptomology. This distinction is critical as treatment for somatization is distinctly different than treatment for mTBI.^35^ Without appropriate identification of somatization, patients cannot be connected with effective interventions. This puts them at high risk for iatrogenic effects from unnecessary medical treatments,^11^ as well as potential worsening by well-meaning clinicians advising typical interventions for mTBI such as rest and symptom avoidance.

Prior authors have raised skepticism about whether symptoms after mTBI represent a true syndrome – a constellation of symptoms that predictably and uniquely co-occur.^36, 37^ If somatization is a major mechanism underlying persistent symptoms, we might expect unclear boundaries between what are typically referred to as ‘post-concussion’ symptoms and other kinds of somatic symptoms, and that the PHQ-15 and the modified 11-item version would have been similarly elevated as compared to the RPQ in patients with prolonged recovery from mTBI. We found that both mTBI-related symptoms and symptoms atypical of mTBI were significantly higher among patients with poor recovery from mTBI when compared to both the control and good recovery groups. However, our results indicate that relative to atypical somatic symptoms, mTBI-related symptoms are more strongly associated with poor outcome. Several explanations are possible. First, somatization may only play a role in a subset of patients in our sample. In a cohort with higher depression and anxiety scores (more typical in patients with continued symptoms and poor recovery), for example, we might expect to see somatization as a more robust variable. Had we therefore not excluded those participants with a prior history of psychiatric problems it is possible that our effect sizes for the atypical symptom scores would be higher. Second, somatization may exacerbate symptoms from the mTBI. Thus ‘typical’ symptoms may be higher due to the combination of both the organic symptoms somatization. Third, knowledge about mTBI and past experience of concussion and its typical symptoms may modify expectations or direct attention (somatic vigilance),^38^ and support symptom misattributions. Somatization, which often coincides with these phenomena, would therefore be more likely to produce typical ‘post-concussion’ symptoms than atypical symptoms (e.g., GI upset) in individuals with more extensive experience and knowledge about concussion.

This study has several limitations. It is comprised of modest sample sizes, and for this study we inventoried symptoms at only a single point in time. As we did not query pre-injury somatization, we cannot be sure somatization scores pre-injury were a risk factor for protracted recovery, or whether protracted recovery led to higher somatization scores. Future research is required in order to determine whether somatoform pathology is a cause or consequence of persistent symptoms after mTBI. Further, we examined somatic symptoms using the PHQ-15, a proxy for examining somatization. While the PHQ-15 has moderate-to-good accuracy for diagnosing somatization, the gold standard for diagnosing somatoform illness is a physician administered structured interview based on the DSM-V. Finally, as our samples were recruited cross-sectionally and immediately post-injury, and we did not take a history of somatic symptoms prior to injury, we were unable to determine whether somatic symptoms were present before, or appeared *de novo* following injury.

In contrast, the greatest strength of this study was that our work occurred in two phases, first as a cross-sectional pilot study followed by a prospectively recruited validation cohort which confirmed the findings of our cross-sectional study. Second, we specifically excluded individuals with diagnosed psychiatric illness, which helped to control for more serious depressive or anxiety symptoms that may influence somatization processes separately from mTBI.

In summary, we present evidence for higher somatic symptoms which are atypical to mTBI in individuals with poor recovery from mTBI, when compared with healthy controls and those with good recovery. While we found higher typical mTBI somatic symptoms in those with poor recovery – as would be anticipated – we also found a significantly higher severity of somatic symptoms which were atypical of mTBI in individuals with poor recovery from mTBI. Though future research is needed, these results provide evidence that somatization may be a significant contributor to persistent symptomology following mTBI and highlight a need to comprehensively assess somatofom pathology as a part of mTBI care.

## Data Availability

Data from this study may be shared upon reasonable request.

## ACKNOWLEDGEMENTS

We thank the CanTBI research assistants for their contributions to the study. Funding for this study was provided by the UBC Neuropsychiatry program research fund, Brain Canada, and Genome BC.

## AUTHOR CONTRIBUTION STATEMENT

JS, KG, and WP conceptualized the study. JS performed the statistical analysis and wrote the initial draft with KG. All authors contributed to the interpretation of the results and the final manuscript.

